# Costs, quality-adjusted life years, and value-of-information of different thresholds for the initiation of invasive ventilation in hypoxemic respiratory failure

**DOI:** 10.1101/2023.03.16.23286754

**Authors:** Christopher J Yarnell, Kali Barrett, Anna Heath, Margaret S. Herridge, Rob Fowler, Lillian Sung, David M Naimark, George Tomlinson

**Author notes:** Corresponding author Christopher Yarnell, Medical-Surgical ICU, 10^th^ floor, 585 University Avenue, Toronto, ON, Canada, M5G 1X5. Contributions Concept: CJY, DN, GT Design: CJY, AH, KB, DN, RAF, LS, GT Data analysis and interpretation: All Data acquisition: CJY Drafting, revising for important intellectual content, final approval, and agreement to be accountable: all.

## Abstract

**Objective:** To estimate costs, quality-adjusted life-years, and the value of undertaking a future randomized controlled trial for different oxygenation thresholds used to initiate invasive ventilation in hypoxemic respiratory failure.

**Design:** Model-based cost-utility estimation with individual-level simulation and value-of-information analysis.

**Setting:** Critical care units.

**Participants:** Adults admitted to critical care receiving non-invasive oxygen.

**Interventions:** We compared four strategies: initiation of invasive ventilation at thresholds of saturation-to-inspired oxygen fraction ratio (SF) < 110, < 98, or < 88, and usual care.

**Main results:** An invasive ventilation initiation threshold of SF < 110, compared to usual care, resulted in more predicted invasive ventilation (62% vs 31%), hospital survival (78.4% vs 75.5%), quality-adjusted life years (QALYs) (8.48 vs 8.34), and lifetime costs (86,700 Canadian dollars (CAD) vs 75,600 CAD). Among the four strategies, threshold SF < 110 had the highest expected net monetary benefit (761,000 CAD), but there was significant uncertainty, because all four strategies had similar probability (range: 23.5% to 27.5%) of having the best net monetary benefit. The expected value to society over the next 10 years of a 400-person randomized trial of oxygenation thresholds was 4.27 billion CAD, and remained high (2.64 billion CAD) in a scenario analysis considering a hypothetical threshold that resulted in less invasive ventilation and similar survival compared to usual care.

**Conclusion:** The preferred threshold to initiate invasive ventilation in hypoxemic respiratory failure is uncertain. It would be highly valuable to society to identify thresholds that, in comparison to usual care, either improve survival or reduce invasive ventilation without reducing survival.

**Key points:** 

**Question:** What are the costs and quality-adjusted life-years associated with different oxygenation thresholds for initiating invasive ventilation, and what is the expected value to society of a randomized controlled trial?

**Findings:** In this health economic evaluation comparing usual care to three different thresholds for initiating invasive ventilation in hypoxemic respiratory failure based on the saturation-to-inspired oxygen fraction ratio (SF), we found that threshold SF < 110 had the highest expected quality-adjusted life-years and net monetary benefit, despite increased predicted invasive ventilation use. However, there was significant residual uncertainty, and the expected value to society of a 400-person randomized trial to compare thresholds for initiating invasive ventilation was greater than 2.5 billion Canadian dollars.

**Meaning:** The preferred threshold to initiate invasive ventilation in hypoxemic respiratory failure is uncertain and further study would be valuable to society.

**Social media summary:** When should we intubate and start invasive ventilation for people with hypoxemic respiratory failure? Our health economic evaluation shows that the preferred threshold is uncertain, but that a clinical trial to determine such a threshold would be immensely valuable to patients and society

## Introduction

Acute hypoxemic respiratory failure comprises at least 10% of admissions to intensive care units (ICUs) worldwide, is associated with ICU mortality of 25-40%, and can cause long-term functional impairment in survivors.(1–4) Invasive ventilation is a potentially life-saving therapy instituted in severe cases, but it can lead to adverse events, including periprocedural cardiac arrest(5,6), ventilator-associated pneumonia, delirium, and ICU-acquired weakness.(7–9) The clinical decision to initiate invasive ventilation is based on expert opinion and clinical experience, with little data available to identify people who truly benefit. However, recent research has suggested that initiating invasive ventilation at less severe hypoxemia in populations with high baseline mortality risk, which would increase the use of invasive ventilation, may improve 28-day survival.(10,11)

The provision of invasive ventilation is resource-intensive.(12–14) Prior efforts to study the cost-effectiveness of invasive ventilation have focused on adjunct therapies such as ventilator management, proning, pulmonary artery catheters, or extracorporeal membrane oxygenation (ECMO).(12,14–18) The resource consequences of adopting different oxygenation thresholds to trigger invasive ventilation have not been estimated. The economic value to society of a randomized trial of thresholds for initiating invasive ventilation is also unknown.

In order to clarify the tradeoffs between short-term mortality, long-term disability, and resource use; and to help quantify the potential economic value of a randomized controlled trial on this topic; we performed a cost-utility and value-of-information analysis comparing usual care to three different oxygenation thresholds for the initiation of invasive ventilation.

## Methods

We performed a model-based cost-utility and value-of-information analysis comparing usual care and three different proposed oxygenation thresholds for initiation of invasive ventilation for people with acute hypoxemic respiratory failure. We adopted the perspective of the publicly funded healthcare payer, a lifetime horizon, and discounted future outcomes by 1.5% annually. We followed the Consolidated Health Economic Evaluation Reporting Standards (checklist in Supplement), and recommendations from the Canadian Agency for Drugs and Technologies in Health.(13,19–21) All analysis code is available at https://doi.org/10.5281/zenodo.7603995.

### Reference case

The reference case was an adult at admission to ICU with non-COVID acute hypoxemic respiratory failure, receiving oxygen via high-flow nasal cannula, non-rebreather mask, or non-invasive positive pressure ventilation.

### Strategies

In the primary scenario we compared four strategies: usual care and initiation of invasive ventilation when saturation-to-inspired oxygen fraction ratio (SF) fell below 110, 98, or 88. Lower SF values indicate more severe hypoxemia. The SF ratio is an attractive choice for threshold definition because it is a simple yet accurate measure of hypoxemia applicable to acute care settings worldwide. It can be measured without blood sampling.(22,23)

Clinically, the three target values correspond to steeper parts of the oxyhemoglobin dissociation curve at high inspired oxygen fractions. Using a non-linear formula to impute arterial-to-inspired oxygen ratios (PF), SF 88, 98, and 110 correspond to PF of at least 55, 68, and 85 mmHg.(24,25) These thresholds have been recently evaluated in an observational study that provides estimates of 28-day invasive ventilation rates and survival for each threshold.(11) We also included a “usual care” strategy, where invasive ventilation occurred at clinical discretion without the use of specified thresholds.

### Model structure and outputs

We built an individual-level simulation model to describe critical illness (Figure 1). We used two-dimensional Monte Carlo simulation to understand how uncertainty affects the model outputs, first sampling model input parameters (such as the probability of hospital survival) 1000 times from their respective distributions (2^nd^ order Monte Carlo), and then simulating the clinical course of 1000 individuals from oxygen therapy through to death (1^st^ order Monte Carlo) for each set of 2^nd^ order samples.

**Figure 1:**
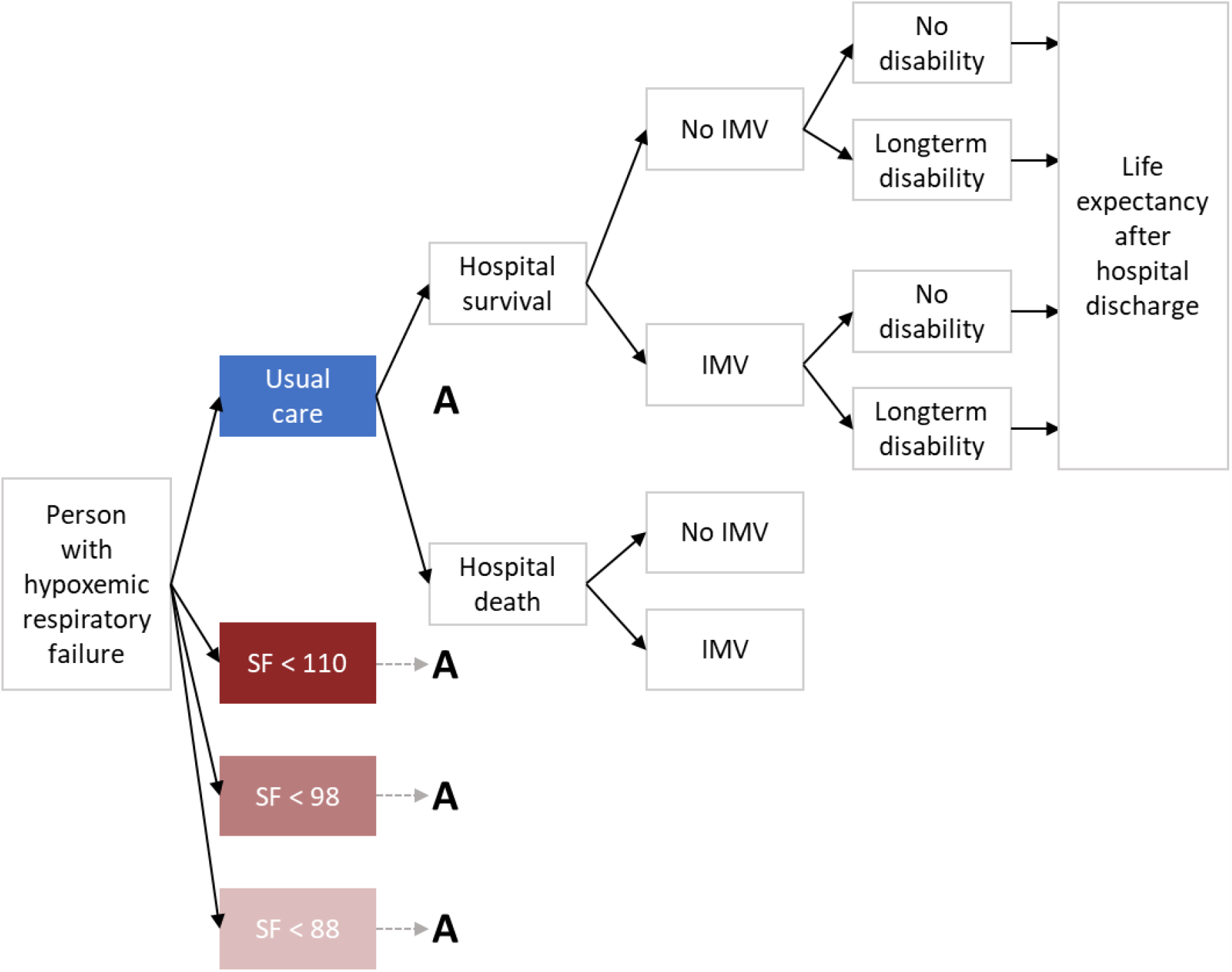
Diagram of health economic model. This diagram shows the model structure. An individual began on the left, at “Person with hypoxemic respiratory failure.” Each individual was simulated through each of the four strategies, with the same tree structure for each threshold (**A**). First, we simulated individuals’ hospital survival, then whether they received invasive ventilation. For survivors, we simulated whether they sustained long-term disability, and finally simulated the remaining life-expectancy after hospital discharge.

Model outputs included clinical outcomes and costs averaged over both 1^st^ and 2^nd^ order iterations. The primary clinical outcome was quality-adjusted life expectancy denominated in years (QALYs). Secondary clinical outcomes were hospital survival, life-years, receipt and duration of invasive ventilation, duration of non-invasive oxygen therapy in ICU, duration of hospital stay after ICU discharge, and incidence of long-term disability. Costs included the cost of intensive care with and without invasive ventilation, ongoing hospitalization after ICU discharge, and ongoing health care during the rest of a survivor’s life post-discharge. Costs were denominated in 2022 Canadian dollars.

### Key assumptions

All people were cared for in an intensive care unit. Invasive ventilation availability for individuals was unaffected by invasive ventilation use in others, approximating usual critical care, as opposed to care during pandemic surges where human and ventilator resources may be scarce.(26–28) We assumed no deviation from thresholds, no re-initiation of invasive ventilation, no ICU readmission, and no utility during hospitalization. Societal willingness-to-pay (WTP) was set at an incremental 100,000 Canadian dollars per QALY gained.(19,29)

### Data sources and parameters

We performed a literature search for each parameter. We favored recent estimates specific to the North American or Northern European context (Tables e2, e3, e4, e5).

### Participant characteristics

The distribution of participant sex and age were based on a cohort of people similar to the reference patient from the Medical Information Mart for Intensive Care version IV (MIMIC-IV) database of 76,540 ICU admissions to an academic hospital in the USA.(30,31)

### Clinical events

For each individual, we first simulated hospital survival according to threshold and then simulated whether invasive ventilation occurred conditional on survival and threshold. The probabilities of invasive ventilation and hospital survival followed a beta distribution with mean event probability based on an observational study of oxygenation thresholds for initiating invasive ventilation.(11) Although the study estimated 28-day outcomes, we used these estimates for hospital outcomes because more comprehensive estimates were not available.

We increased the standard deviation in the estimates from the observational study 5-fold from 0.02 to 0.1, because the data came from a single center. This was analogous to a 25-fold decrease in sample size, meaning that we treated the estimate from the 3,357-person observational study as if it provided as much information as a randomized trial enrolling approximately 134 people.(32)

Hospital survivors had the remainder of their lifespan drawn from a Weibull distribution based on a cohort of intensive care unit survivors with 5 years of follow-up, in order to incorporate the increased time-varying risk of death faced by survivors of critical illness (Figure e1).(4)

We used time-to-event models to simulate durations of invasive ventilation, ICU stay while not invasively ventilated, and hospital stay after ICU discharge. We fit Weibull curves to the same cohort that was used to derive the parameter estimates for invasive ventilation and hospital survival. For the model describing duration of invasive ventilation, we included age, sex, and survival as covariates; for duration of oxygen therapy and hospital stay we used the same covariates with the addition of a binary covariate for receipt of invasive ventilation.(33,34)

The probability of long-term disability was based on a cohort of mechanical ventilation survivors, which divided people into disability risk groups according to age, duration of ventilation, and length of ICU stay.(35) In our model, survivors of mechanical ventilation were divided into the same groups. The probability of long-term disability followed a beta distribution, with mean probability within each group equal to the observed probability of requiring assistance with bathing at 12-months.

### Utilities

We used population-based estimates to estimate utility in survivors.(36) Survivors with long-term disability incurred an ongoing disutility with mean 0.15, which was subtracted from their population-based utility. This was based on prior work showing persistent decreased utility in critical illness survivors out to at least five years.(4)

### Costs

Costs accumulated during hospitalization and any subsequent recovery. To estimate daily ICU cost while ventilated, ICU cost while not ventilated, and ward cost after ICU discharge, we combined data from a study based in ICUs in Ottawa, Canada and a national survey of Canadian ICUs.(37,38) We estimated mean annual costs for hospital survivors with data from a cohort of acute respiratory distress syndrome survivors (3) and administrative data from Ontario, Canada.(39) All costs were converted to CAD, adjusted for inflation, and reported in 2022 CAD.(40) Our model did not capture costs not covered by provincial health insurance, for example, dental care, or outpatient medications for those under 65 years. We did not incorporate economic losses due to underemployment of survivors or care partners.

### Analysis

We compared strategies using the net monetary benefit, defined as *QALYS x WTP – costs*. We reported the incremental cost-utility ratio for non-dominated strategies relative to usual care. We built the model in TreeAge Pro (v2021 R2) and analyzed outputs in R.(34,41) We reported means and 95% credible intervals (CrI). We showed, for each strategy and at WTP ranging from 0 to 200,000 CAD, the probability of having the highest net monetary benefit.

### Value-of-information analysis

Uncertainty in model inputs leads to uncertainty in model outputs. Broad adoption of an invasive ventilation initiation threshold based on the average model output may be the best decision using current knowledge. However, collecting additional information through a randomized controlled trial may reveal that the current best decision is incorrect.

The economic consequence of an incorrect decision is referred to as the opportunity-cost. Value-of-information analysis quantifies the opportunity-cost of deciding now, with imperfect information, compared to an alternative decision once more information is available.(42) We can compare the net monetary benefits of a decision made with perfect information (no uncertainty in model parameters) and a decision made with current, imperfect information. The difference is termed the expected value of perfect information (EVPI). Complete certainty for all input parameters is unrealistic, so we also consider the value of perfect information in a subset of input parameters, called the expected value of partial perfect information (EVPPI). However, a randomized trial cannot provide complete certainty, at best it can reduce uncertainty. The economic value of this reduction is the expected value of sample information (EVSI).(43) The EVSI can be calculated for different sample sizes.

We calculated EVPI, EVPPI, and EVSI for a proposed randomized controlled trial. We used the Sheffield value-of-information application for EVPI and EVPPI, and non-parametric regression for EVSI.(44–46) For EVPPI, we included the parameters for invasive ventilation rate and hospital survival probability according to initiation threshold. We calculated the EVSI for randomized trials with sample sizes ranging from 100 to 3200 participants. We did not evaluate EVSI for trials with less than 100 participants, which would be too small to effect any change in clinical practice. We combined these results with the annual number of Canadian ICU admissions (38) and incidence of acute respiratory failure (1) to calculate population EVSI.

### Scenario analysis

We considered two scenarios. The first scenario is described above, where there exists a threshold for the initiation of invasive ventilation associated with increased use of invasive ventilation and improved survival compared to usual care. In the second scenario, we compared a hypothetical threshold to usual care, where the hypothetical threshold had used less invasive ventilation with equivalent survival compared to usual care. We repeated the analysis to assess the expected value of sample information if we could identify thresholds that reduced the use of invasive ventilation while maintaining equivalent survival.

## Results

### Clinical outcomes

Invasive ventilation ranged from 20.7% with threshold SF < 88 to 62.1% with threshold SF < 110 (Table 1). Hospital survival ranged from 73.8% with threshold SF < 88 to 78.4% with threshold SF < 110. Among additional hospital outcomes, the mean durations of ICU admission while not ventilated, ICU admission while ventilated, and ward stay after ICU discharge were similar across strategies. Long-term disability varied from 4.6% with threshold SF < 88 to 11.4% with threshold SF < 110.

**Table:**
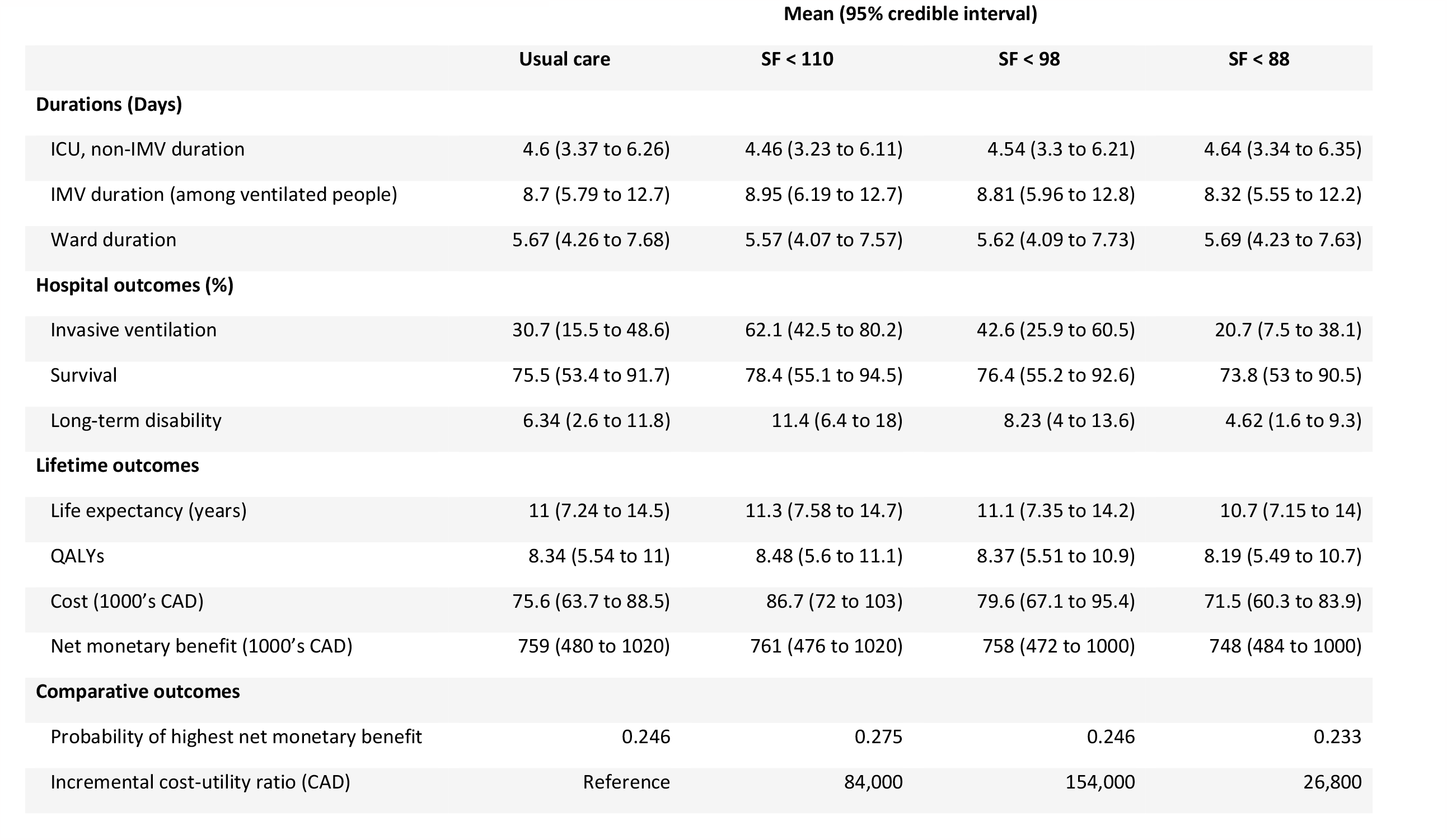
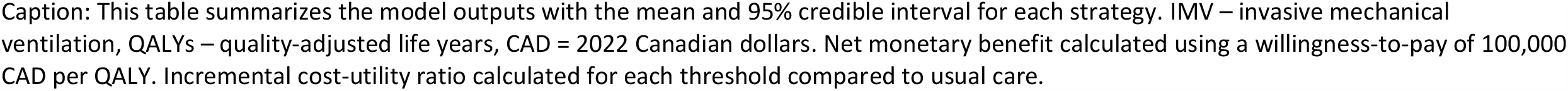
Model outputs, primary analysis.

**Table: Model outputs, primary analysis**
|  | Mean (95% credible interval) |  |  |  |
| --- | --- | --- | --- | --- |
|  | Usual care | SF < 110 | SF < 98 | SF < 88 |
| **Durations (Days)** |  |  |  |  |
| ICU, non-IMV duration | 4.6 (3.37 to 6.26) | 4.46 (3.23 to 6.11) | 4.54 (3.3 to 6.21) | 4.64 (3.34 to 6.35) |
| IMV duration (among ventilated people) | 8.7 (5.79 to 12.7) | 8.95 (6.19 to 12.7) | 8.81 (5.96 to 12.8) | 8.32 (5.55 to 12.2) |
| Ward duration | 5.67 (4.26 to 7.68) | 5.57 (4.07 to 7.57) | 5.62 (4.09 to 7.73) | 5.69 (4.23 to 7.63) |
| **Hospital outcomes (%)** |  |  |  |  |
| Invasive ventilation | 30.7 (15.5 to 48.6) | 62.1 (42.5 to 80.2) | 42.6 (25.9 to 60.5) | 20.7 (7.5 to 38.1) |
| Survival | 75.5 (53.4 to 91.7) | 78.4 (55.1 to 94.5) | 76.4 (55.2 to 92.6) | 73.8 (53 to 90.5) |
| Long-term disability | 6.34 (2.6 to 11.8) | 11.4 (6.4 to 18) | 8.23 (4 to 13.6) | 4.62 (1.6 to 9.3) |
| **Lifetime outcomes** |  |  |  |  |
| Life expectancy (years) | 11 (7.24 to 14.5) | 11.3 (7.58 to 14.7) | 11.1 (7.35 to 14.2) | 10.7 (7.15 to 14) |
| QALYs | 8.34 (5.54 to 11) | 8.48 (5.6 to 11.1) | 8.37 (5.51 to 10.9) | 8.19 (5.49 to 10.7) |
| Cost (1000's CAD) | 75.6 (63.7 to 88.5) | 86.7 (72 to 103) | 79.6 (67.1 to 95.4) | 71.5 (60.3 to 83.9) |
| Net monetary benefit (1000's CAD) | 759 (480 to 1020) | 761 (476 to 1020) | 758 (472 to 1000) | 748 (484 to 1000) |
| **Comparative outcomes** |  |  |  |  |
| Probability of highest net monetary benefit | 0.246 | 0.275 | 0.246 | 0.233 |
| Incremental cost-utility ratio (CAD) | Reference | 84,000 | 154,000 | 26,800 |

Caption: This table summarizes the model outputs with the mean and 95% credible interval for each strategy. IMV – invasive mechanical ventilation, QALYs – quality-adjusted life years, CAD = 2022 Canadian dollars. Net monetary benefit calculated using a willingness-to-pay of 100,000 CAD per QALY. Incremental cost-utility ratio calculated for each threshold compared to usual care.

There were small differences in life expectancy and QALYs across thresholds, and larger relative differences in costs. The lowest mean lifetime costs were 71,500 CAD (SF < 88), while the highest mean costs were 86,700 CAD (SF < 110).

### Comparing strategies

At the prespecified willingness-to-pay of 100,000 CAD, mean net monetary benefit ranged from 748,000 CAD (SF < 88) to 761,000 CAD (SF < 110). Among the four strategies, threshold SF < 110 was most likely to have the highest net monetary benefit. However, each threshold’s probability of having the highest net monetary benefit was similar, indicating significant residual uncertainty in the model outputs. Figure 2 shows the similarity between the probabilities of each threshold having the highest net monetary benefit. The incremental cost-utility ratio for each threshold when compared to usual care was 84,000 CAD per QALY for SF < 110, 154,000 CAD per QALY for SF < 98, and 27,500 per QALY for SF < 88.

**Figure 2:**
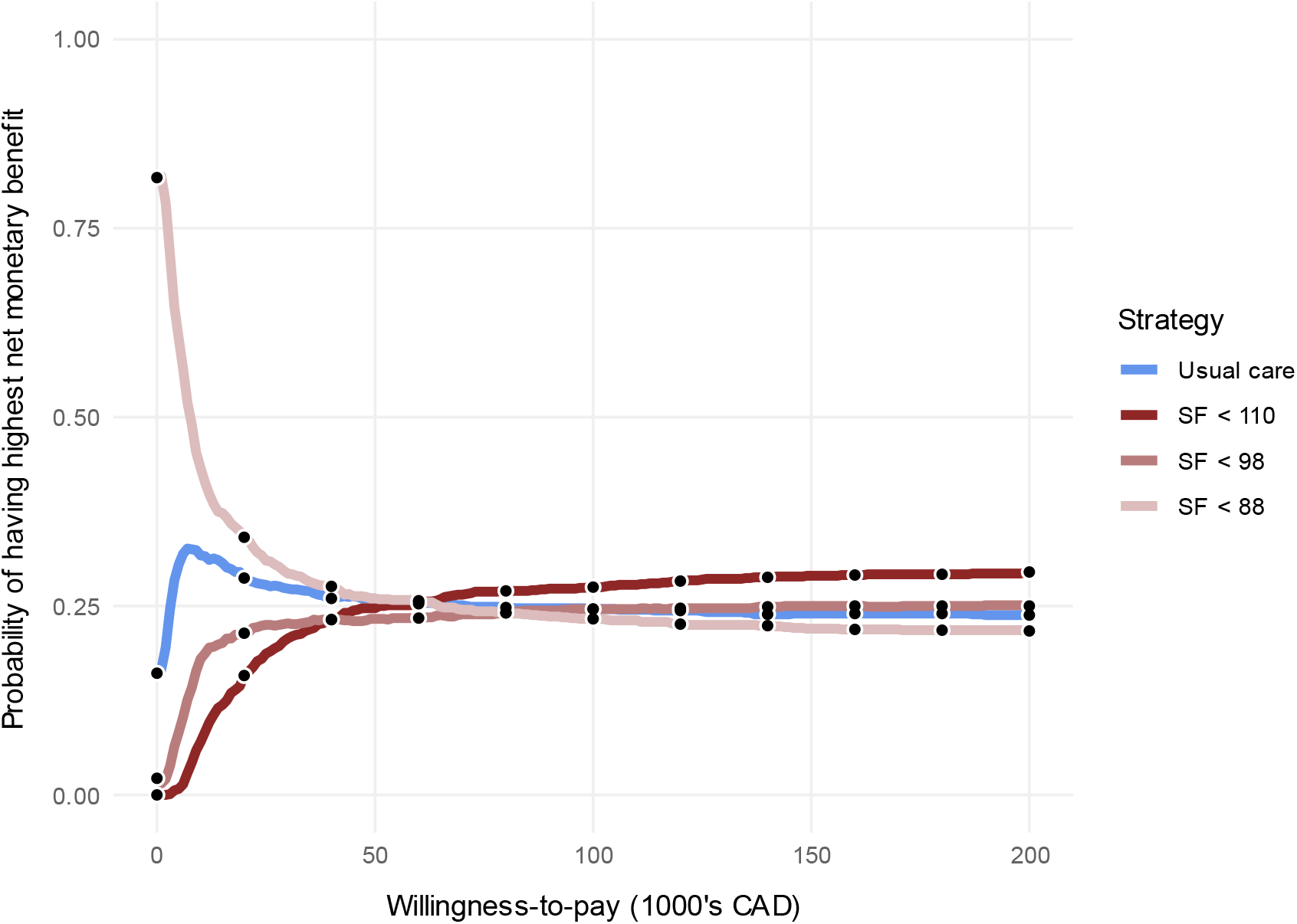
Cost-effectiveness acceptability curve. This figure shows the proportion of iterations that each strategy has the highest net monetary benefit versus willingness to pay. SF < 88 is the optimal choice until a willingness-to-pay of approximately 50,000 CAD, after which there is uncertainty until SF < 110 emerges as the optimal choice for willingness-to-pay values greater than 65,000 CAD.

### Value-of-information

At a willingness-to-pay of 100,000 CAD, the EVPI per person was 104,700 CAD. EVPPI for the randomized trial group of parameters was 100,300 CAD and captured almost all of the value of perfect information. A randomized trial of 400 patients had EVSI of 91,333 CAD per person.

Using a national annual incidence of 5,000 eligible people per year (38), a discount rate of 1.5%, and a 10-year horizon over which the information will be useful, population EVSI for a randomized controlled trial of 400 participants was 4.27 billion CAD.

### Scenario analysis

Recall that the second scenario considered a hypothetical threshold which used less invasive ventilation and achieved similar survival compared to usual care. The probability of invasive ventilation was reduced from 28% with usual care to 20% with the hypothetical threshold, without a difference in survival (75%) (Table e6). The hypothetical threshold was most likely to have the highest net monetary benefit up to a willingness-to-pay value of 100,000 CAD, after which the net monetary benefit was equal (Figure e2). Per-person EVPI was 62,900 CAD and per-person EVPPI of a randomized controlled trial was 60,200 CAD. EVSI of a randomized controlled trial with 400 participants was 56,500 CAD, corresponding to a 10-year population EVSI of 2.64 billion CAD. In Figure 3 we show the population EVSI associated with randomized controlled trials of varying sample sizes for both scenarios.

**Figure 3:**
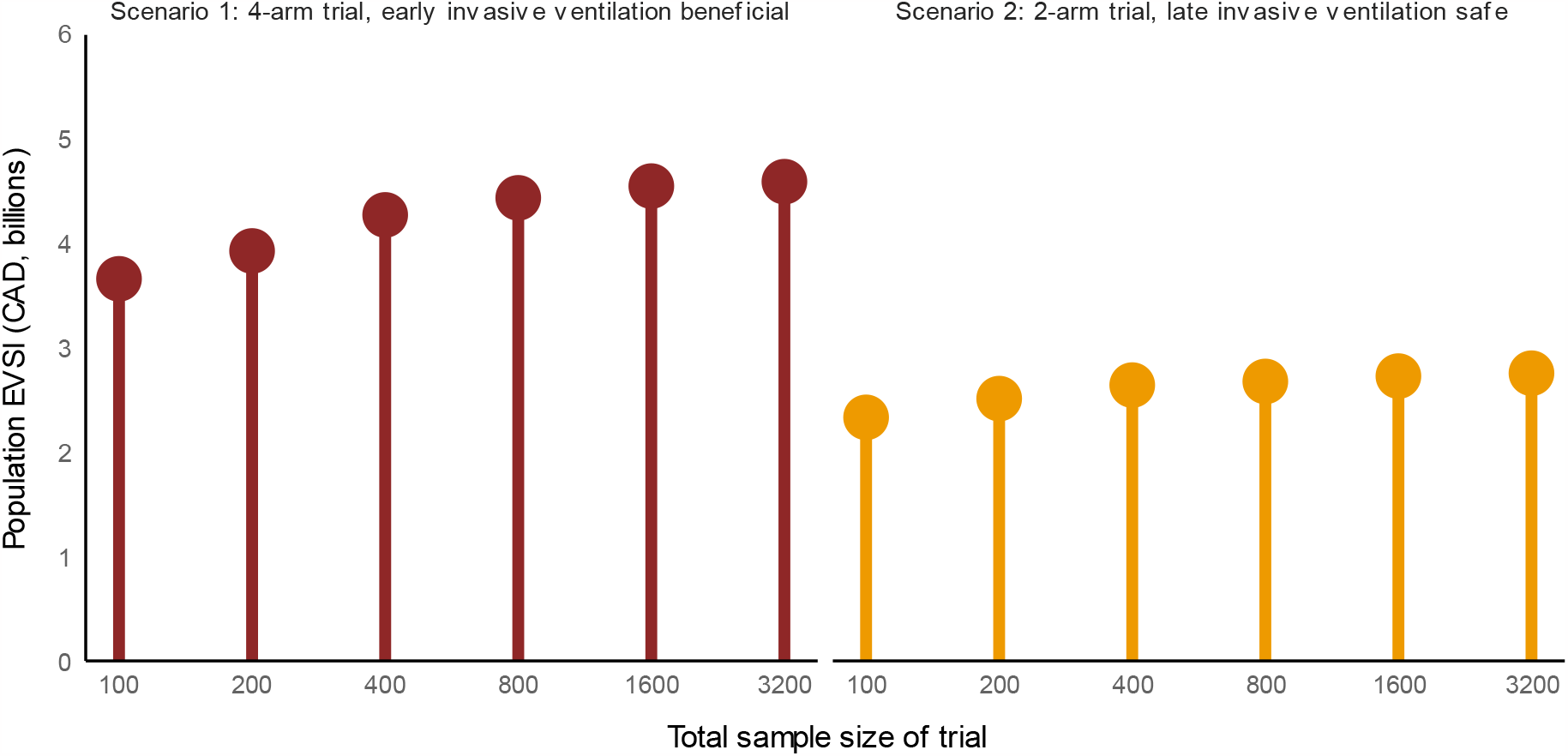
Population expected value of a randomized trial by sample size. This figure shows the population expected value of sample information (EVSI, y-axis) for randomized trials of different total sizes (x-axis, logarithmic scale) for both scenario 1 (left) and scenario 2(right). The expected value of sample information captures the value of the additional information gleaned from a study of a particular size. The value comes from an improved probability of correctly identifying the most cost-effective strategy. The population EVSI is derived by multiplying the per-person EVSI by the annual incidence of eligible people in Canada (5000) and the duration of information relevance (10 years), discounting future benefits by 1.5% per year. For these values of the EVSI we used a willingness-to-pay of 100,000 CAD. Note that these values only describe value that accrues to the Canadian population. EVSI = expected value of sample information, CAD = 2022 Canadian dollars.

## Discussion

In this health economic evaluation comparing usual care to three different oxygenation thresholds for initiating invasive ventilation in hypoxemic respiratory failure, we found that the threshold SF < 110 had the highest expected net monetary benefit, despite increased predicted resource use and long-term disability. However, the probabilities of each threshold having the highest net monetary benefit among the four strategies were similar, meaning that there was significant residual uncertainty. The expected value to Canadian society of a 400-person randomized controlled trial, based on the potential improvement in QALYs and a willingness-to-pay of 100,000 CAD per QALY, was 4.27 billion dollars. The expected value of such a trial remained high (2.64 billion CAD), even if the best alternative to usual care was a threshold with lower invasive ventilation and equivalent mortality. These findings suggest that there is uncertainty about the preferred threshold for initiating invasive ventilation, and that further research on this question would be immensely valuable for society.

The findings build on prior work by illustrating the net impact of survival, ventilator resource use, and disability on cost-effectiveness, in the setting where increased invasive ventilation is associated with improved survival.(47) The SF < 110 threshold had a higher net monetary benefit than usual care, after combining the QALY impact of additional survivors (3% absolute increase), increased invasive ventilation use (31% absolute increase), and increased long-term disability (5% absolute increase). The increase in predicted long-term disability resulted from increased predicted use of invasive ventilation, which shifted patients to higher long-term disability risk groups. Prior work has investigated the mortality associated with particular thresholds for invasive ventilation but has not incorporated functional outcomes or costs (10,11,48). It is somewhat reassuring that small improvements in survival associated with increased use of invasive ventilation can be cost-effective. However, individuals may vary in the extent to which they value a life affected by weakness, dependence, and post-traumatic stress disorder.(3) Further research needs to clarify what increase in long-term disability risk is acceptable to people in this situation.

These results also offer insights applicable to respiratory pandemics or resource-constrained settings, despite using data from before the coronavirus-19 pandemic (COVID-19) and assuming critical care capability typical of developed nations. Settings where the availability of technology, personnel, or funding is limited could be approximated by a lower willingness-to-pay, and our results show that as willingness-to-pay declines, the advantages of the most severe hypoxemia threshold (SF < 88) increase relative to other strategies. Our model did not consider ICU capacity as a constraint, and we did not consider the impact of SF thresholds on population-level ICU bed requirements. Future modelling considering health system capacity requirements at different SF thresholds needs to be considered.

The expected value of a randomized trial over 10 years on this topic was more than 2.5 billion CAD and far exceeded the cost of the proposed trial, in both scenarios. The median cost of a clinical trial with more than 1000 participants testing a novel pharmaceutical agent is approximately 70 million CAD.(49) Our EVSIs could be overestimated because the value captured in the EVSI assumes that all clinicians will incorporate the study findings into their decision-making.(18,50) However, our population EVSIs could also be underestimated because they correspond only to Canada, even though similar benefits could accrue to eligible people all over the world.

This work has additional limitations. It is a model-based evaluation and may not incorporate enough nuance from clinical practice to be useful. Our results depend on many decisions made in the course of model construction; different decisions may have led to different results.(51) We did not include patient engagement. Our results are most applicable to a Canadian context. The thresholds rely on oxygen saturation, which can be overestimated when measured with a pulse oximeter in people with darker skin pigment.(52) Some of the costing data used was from more than 10 years ago and may not reflect the costs of high-flow nasal cannula which have become commonly used for non-intubated patients with hypoxemic respiratory failure.

However, a prior health economic analysis in the UK found that high-flow nasal cannula use, compared to standard oxygen or non-invasive ventilation, was likely to be cost saving.(53) The costing data from a prior ARDS cohort is also limited because it focused on a younger group of patients.(3) Our results are also limited by parameter validity. In particular, the parameters did not incorporate threshold-related mortality beyond 28 days, because of data limitations. We also do not have information on whether the probability of long-term disability varies by threshold choice, beyond the extent to which it is affected by the use of invasive ventilation.

We included parameter uncertainty to address these limitations.

## Conclusion

The preferred threshold to initiate invasive ventilation in hypoxemic respiratory failure is uncertain. It would be highly valuable to society to identify thresholds that, in comparison to usual care, either improve survival or reduce invasive ventilation without reducing survival.

## Supporting information

Electronic supplement

## Data Availability

All data for this study are either published in scientific journals or publicly available through Physionet (Medical Information Mart for Intensive Care version IV, https://physionet.org/content/mimiciv/2.2/).

## Acknowledgements

We thank the following for helpful comments: Allan Detsky.

