## Supplementary material for "Costs, quality-adjusted life years, and value-of-information of different thresholds for the initiation of invasive ventilation in hypoxemic respiratory failure": Electronic supplement

### Cost-utility and value of information analysis of oxygenation thresholds for invasive ventilation in hypoxemic respiratory failure

Yarnell, Christopher J MD; ORCID 0000-0001-5657-9398

Barrett, Kali

Heath, Anna

Herridge, Margaret

Fowler, Rob

Sung, Lillian

Naimark, David

Tomlinson, George

2023-03-02

### 1 CHEERS checklist

| Topic | No. | Item | Location where item is reported |
|---|---|---|---|
| **Title** | | | |
| | 1 | Identify the study as an economic evaluation and specify the interventions being compared. | Title, Abstract, Methods |
| **Abstract** | | | |
| | 2 | Provide a structured summary that highlights context, key methods, results, and alternative analyses. | Abstract (page 4) |
| **Introduction** | | | |
| **Background and objectives** | 3 | Give the context for the study, the study question, and its practical relevance for decision making in policy or practice. | Introduction |
| **Methods** | | | |
| **Health economic analysis plan** | 4 | Indicate whether a health economic analysis plan was developed and where available. | Methods |
| **Study population** | 5 | Describe characteristics of the study population (such as age range, demographics, socioeconomic, or clinical characteristics). | Methods |
| **Setting and location** | 6 | Provide relevant contextual information that may influence findings. | Methods, Paragraphs 3 & 4 |
| **Comparators** | 7 | Describe the interventions or strategies being compared and why chosen. | Methods, paragraphs 3 & 4 |

| Topic | No. | Item | Location where item is reported |
|---|---|---|---|
| **Perspective** | 8 | State the perspective(s) adopted by the study and why chosen. | Methods, paragraph 1 |
| **Time horizon** | 9 | State the time horizon for the study and why appropriate. | Methods, paragraph 1 |
| **Discount rate** | 10 | Report the discount rate(s) and reason chosen. | Methods, paragraph 1 |
| **Selection of outcomes** | 11 | Describe what outcomes were used as the measure(s) of benefit(s) and harm(s). | Methods, "Model structure and outputs" paragraph 2 |
| **Measurement of outcomes** | 12 | Describe how outcomes used to capture benefit(s) and harm(s) were measured. | Methods, "Model structure and outputs" paragraph 2 |
| **Valuation of outcomes** | 13 | Describe the population and methods used to measure and value outcomes. | Methods, "Model structure and outputs" paragraph 2 |
| **Measurement and valuation of resources and costs** | 14 | Describe how costs were valued. | Methods, "Model structure and outputs" paragraph 2 |
| **Currency, price date, and conversion** | 15 | Report the dates of the estimated resource quantities and unit costs, plus the currency and year of conversion. | Methods, "Model structure and outputs" paragraph 2, and Supplement, section 3.2 |
| **Rationale and description of model** | 16 | If modelling is used, describe in detail and why used. Report if the model is publicly available and where it can be accessed. | Methods, first paragraph, and "Model structure and outputs", and Figure 1 |

| Topic | No. | Item | Location where item is reported |
|---|---|---|---|
| **Analytics and assumptions** | 17 | Describe any methods for analysing or statistically transforming data, any extrapolation methods, and approaches for validating any model used. | Methods , Last paragraph and Supplement |
| **Characterising heterogeneity** | 18 | Describe any methods used for estimating how the results of the study vary for subgroups. | None used |
| **Characterising distributional effects** | 19 | Describe how impacts are distributed across different individuals or adjustments made to reflect priority populations. | Not incorporated |
| **Characterising uncertainty** | 20 | Describe methods to characterise any sources of uncertainty in the analysis. | Methods, "Model structure and outputs" |
| **Approach to engagement with patients and others affected by the study** | 21 | Describe any approaches to engage patients or service recipients, the general public, communities, or stakeholders (such as clinicians or payers) in the design of the study. | Not done |
| **Results** | | | |
| **Study parameters** | 22 | Report all analytic inputs (such as values, ranges, references) including uncertainty or distributional assumptions. | Supplementary tables e1, e2, e3, e4 |
| **Summary of main results** | 23 | Report the mean values for the main categories of costs and outcomes of interest and summarise them in the most appropriate overall measure. | Results |

| Topic | No. | Item | Location where item is reported |
|---|---|---|---|
| **Effect of uncertainty** | 24 | Describe how uncertainty about analytic judgments, inputs, or projections affect findings. Report the effect of choice of discount rate and time horizon, if applicable. | Results |
| **Effect of engagement with patients and others affected by the study** | 25 | Report on any difference patient/service recipient, general public, community, or stakeholder involvement made to the approach or findings of the study | Not reported |
| **Discussion** | | | |
| **Study findings, limitations, generalisability, and current knowledge** | 26 | Report key findings, limitations, ethical or equity considerations not captured, and how these could affect patients, policy, or practice. | Discussion |
| **Other relevant information** | | | |
| **Source of funding** | 27 | Describe how the study was funded and any role of the funder in the identification, design, conduct, and reporting of the analysis | End of manuscript |
| **Conflicts of interest** | 28 | Report authors conflicts of interest according to journal or International Committee of Medical Journal Editors requirements. | End of manuscript |

*From:* Husereau D, Drummond M, Augustovski F, et al. Consolidated Health Economic Evaluation Reporting Standards 2022 (CHEERS 2022) Explanation and Elaboration: A Report of the ISPOR CHEERS II Good Practices Task Force. Value Health 2022;25. doi:10.1016/j.jval.2021.10.008

#### 2 Parameters for invasive ventilation and survival

The parameters for invasive ventilation and survival according to threshold were drawn from a target trial emulation of oxygenation thresholds for invasive ventilation conducted in two cohorts.(1)

The primary cohort studied in this target trial was drawn from the Medical Information Mart for Intensive Care (MIMIC) version IV database.(2,3) This database uses information from patients cared for in intensive care units at the Beth Israel Deaconess Medical Center in Boston, USA between 2009 and 2019. In the primary analysis from this target trial emulation, including 3,357 patients, the threshold SF < 110 was associated with the best 28-day survival.

The sensitivity analysis uses parameters from the secondary analysis in this target trial emulation, which were derived from the AmsterdamUMCdb database.(4,5) This database is based on patients cared for at the Amsterdam University Medical Centre in Amsterdam, Netherlands, between 2003 and 2016. In the secondary analysis from this target trial emulation, including 1,279 patients, the threshold SF < 88 was associated with the best 28-day survival.

The risk of mortality was overall lower in the cohort of non-intubated patients from Amsterdam. The final conclusion of the study was that for patients at higher baseline risk of mortality, such as those in the MIMIC-IV cohort of non-intubated patients, choosing lower hypoxemia severity thresholds would lead to higher 28-day survival, while for patients at lower baseline risk of mortality, such as those in the AmsterdamUMCdb cohort of non-intubated patients, choosing higher hypoxemia severity thresholds would lead to higher 28-day survival.

#### 2.1 Table e1 – Probabilities of survival and invasive ventilation by oxygenation threshold for invasive ventilation, AmsterdamUMCdb cohort

| Threshold | | Invasive ventilation (28-day) | |
|---|---|---|---|
| | Survival (28 day) | Survivors | Deceased |
| Usual care | 0.86 | 0.44 | 0.57 |
| SF < 110 | 0.85 | 0.49 | 0.64 |
| SF < 98 | 0.87 | 0.26 | 0.36 |
| SF < 88 | 0.87 | 0.18 | 0.24 |

Table caption: This table shows the probability of 28-day clinical events by threshold, based on the target trial emulation by Yarnell et al 2022.(6) All parameters were implemented in the model as beta distributions with the above mean and a standard deviation of 0.1, using the alternative parameterization (mean and standard deviation) of the beta distribution.

### 3 Parameter value tables

#### 3.1 Table e2 – Probabilities of survival and invasive ventilation by oxygenation threshold for invasive ventilation

| Threshold | | Invasive ventilation | |
|---|---|---|---|
| | Hospital survival | Survivors | Deceased |
| Usual care | 0.75 | 0.34 | 0.25 |
| SF < 110 | 0.78 | 0.74 | 0.64 |
| SF < 98 | 0.76 | 0.49 | 0.40 |
| SF < 88 | 0.74 | 0.21 | 0.16 |

Table caption: This table shows the probability of clinical events by threshold, based on the target trial emulation by Yarnell et al 2022.(1)

#### 3.2 Table e3 – Costs during hospitalization

| Day | ICU, on invasive ventilation | ICU, not on invasive ventilation | Ward (after ICU discharge) |
|---|---|---|---|
| 1 | 6415 | 4009 | 2264 |
| 2 | 4567 | 2855 | 1883 |
| 3 | 4140 | 2588 | 1924 |
| 4 | 4037 | 2523 | 1751 |
| 5+ | 3925 | 2453 | 1533 |

Table caption: This table shows the mean cost (in 2022 Canadian dollars) of each successive day of care in an ICU on invasive ventilation, in an ICU not on invasive ventilation, or on the ward after ICU discharge, based on Evans et. al 2018 and Kaier et al.(7,8) Costs were adjusted to 2022 Canadian dollars by first converting to Canadian dollars at the currency conversion rate based on the time of costing data in the study (if costs were not in Canadian dollars), and then adjusting for inflation using the Canadian Consumer Price Index.(9)

#### 3.3 Table e4: Additional parameters

| Type | Name | Distribution | Detail | Values | Source |
|---|---|---|---|---|---|
| **Probability** | Long-term disability | Beta | IMV duration < 7 | mean = 0.05, sd = 0.1 | Expert opinion |
| | | Beta | IMV duration ≥ 7 AND LOS < 14 AND age < 42 | mean = 0.15, sd = 0.15 | Herridge 2016 |
| | | Beta | (IMV duration ≥ 7) AND [(LOS < 14 AND age ≥ 42) OR (LOS ≥ 14 AND age < 45)] | mean = 0.25, sd = 0.2 | Herridge 2016 |
| | | Beta | IMV duration ≥ 7 AND LOS ≥ 14 AND age 45-66 | mean = 0.40, sd = 0.2 | Herridge 2016 |
| | | Beta | Duration of IMV ≥ 7 AND LOS ≥14 AND age >= 66 | mean = 0.60, sd = 0.2 | Herridge 2016 |
| **Costs** | Recovery after hospital discharge | Gamma | Year 1 | mean = 29,595, sd = 7,746 | Herridge 2016 |
| | | Gamma | Year 2 | mean = 13,113, sd = 5,121 | Herridge 2016 |
| | | Gamma | Year 3 | mean = 8,043, sd = 4,011 | Herridge 2016 |
| | | Gamma | Year 4 | mean = 7,627, sd = 3906 | Herridge 2016 |
| | | Gamma | Year 5 | mean = 7,384, sd = 3843 | Herridge 2016 |
| | | Mean – Normal<br>Individual - Gamma | Year 6+, age 18-64 | Mean – mean = 3469, sd = 1000<br>Individual – mean from above, sd = 2000 | Wodchis 2016 |
| | | Mean – Normal<br>Individual - Gamma | Year 6+, age 65+ | Mean – mean = 29911, sd = 5000<br>Individual – mean from above, sd = 5000 | Wodchis 2016 |
| **Utility** | Recovery | Deterministic | | Varies by age and sex | Guertin 2018 |
| | Long-term disability penalty | PERT | annual penalty | mean = 0.15, min = 0.05, max = 0.4 | Cuthbertson 2010 |
| **Lifespan** | Death from recovered state | Weibull | | Shape: mean = 0.4602, sd = 0.0376<br>Scale: mean = 16.711, sd = 3.8 | Life tables 2020 |
| | Hazard ratio for age >= 64 | Log-normal | | Mean = log(2.09), sd = 0.3 | Cuthbertson 2010 |
| **Covariates** | Age | Normal | | mean = 65, sd = 15 | MIMIC-IV |
| | Sex | Beta | Proportion female | mean = 0.45, sd = 0.05 | MIMIC-IV |
| **Discount** | Annual | Deterministic | | 1.50% | CADTH |

Table caption: This table shows the remaining parameters not contained in Tables 1a and 1b, divided by type (left column). IMV = invasive mechanical ventilation, LOS = intensive care unit length of stay, sd = standard deviation, PERT = convex probability distribution parameterized by mean, minimum, and maximum, MIMIC-IV = Medical Information Mart for Intensive Care version IV, CADTH = Canadian Agency for Drugs and Technology in Health

### 4 Additional parameter details

Here we include additional information on relevant parameters.

#### 4.1 Cost of daily care in ICU, ventilated and non-ventilated

We combined multiple sources to estimate the average cost of a day in ICU while ventilated and the average cost of a day in ICU while not ventilated. Prior work from the Netherlands in 2006 suggested an incremental cost of invasive ventilation that converts to 3,539$, in 2022 CAD.(10) However, this is likely high, given that a Canadian Institutes of Health Information report from 2016 reported the average cost of a day in the intensive care unit was 4,301$ in 2022 CAD.(11) The costing data most applicable to this study comes from a 2018 study out of Ottawa, Canada, that broke down ICU costs into fixed and variable components for each of days 1, 2, 3, 4, and 5+ of ICU admission. The variable portion of the daily ICU costs was approximately 85% of the total cost, with fixed costs making up the remaining 50%. This study also included the costs of a hospital ward stay (days 1, 2, 3, 4, 5+) after ICU discharge. Unfortunately, the Ottawa study did not have a breakdown of costs according to whether or not a patient was receiving invasive ventilation (we contacted the author who confirmed this).

We used information from other studies to estimate the differential cost between a ventilated and non-ventilated day in ICU. A German study using data from 2013 found that the daily costs for ventilated patients were 60% higher than costs for non-ventilated patients.(8) An American study from the early 2000s found a similar relative difference.(12) Given that care of a non-ventilated ICU patient with respiratory failure is more similar to care of a ward patient (most likely high-flow nasal cannula use, less likely non-invasive ventilation use) than a ventilated ICU patient, we set the cost of a day in ICU on non-invasive oxygen therapy to be 5/8 of the cost of the full estimated cost of a day in ICU from the Ottawa study.

We also assumed that the average costs on day 1 would be higher than the average costs on day 2 and so on, similar to prior research. Out of concern that enforcing the order of average costs would unduly suppress the variance, we doubled the standard deviation of the cost of the first day of invasive ventilation relative to the standard deviation used for all other daily costs.

#### 4.2 Life expectancy among survivors

ICU survivors face increased risk of death up to at least 5 years after hospital discharge compared to the general population.(13–21) In Cuthbertson et al (2010), they enrolled 300 consecutive consenting patients discharged from a mixed medical/surgical ICU in the United Kingdom. The ICU mortality rate at that time was 24%. The survivors had a median age of 61 years. Compared to an age- and sex-matched UK survival curve, the ICU survivor cohort had decreased survival at months 12 (74% vs 98%), 24 (69% vs 96%), and 60 (57% vs 90%) (extracted from Kaplan Meier curves).(22) This suggests an increased hazard of mortality for ICU survivors that remains elevated for at least 5 years, with the highest hazard occurring in the first 12 months and the hazard decreasing in time after. Factors associated with mortality included age, duration of ICU stay, APACHE II score, and premorbid physical and mental functioning.

We digitally extracted the data from the curve of Cuthbertson et al.(22) We fit a Weibull model to this curve. The shape parameter was 0.4602 (95% confidence interval 0.3922 to 0.5401, standard error 0.0376), and the scale parameter was 16.711 (95% confidence interval 10.7 to 26.1, standard error 3.8, using a time unit of years). Figure e1 shows that the Weibull curve does a reasonable job of describing the time-varying hazard. In our model, we also used the hazard ratio for age less than 64 derived by Cuthbertson, assuming that 50% of patients had an age less than 64. Note that we capped life expectancy for all patients at 100 years of total life duration.

#### 4.3 Figure e1: Model for post-discharge survival

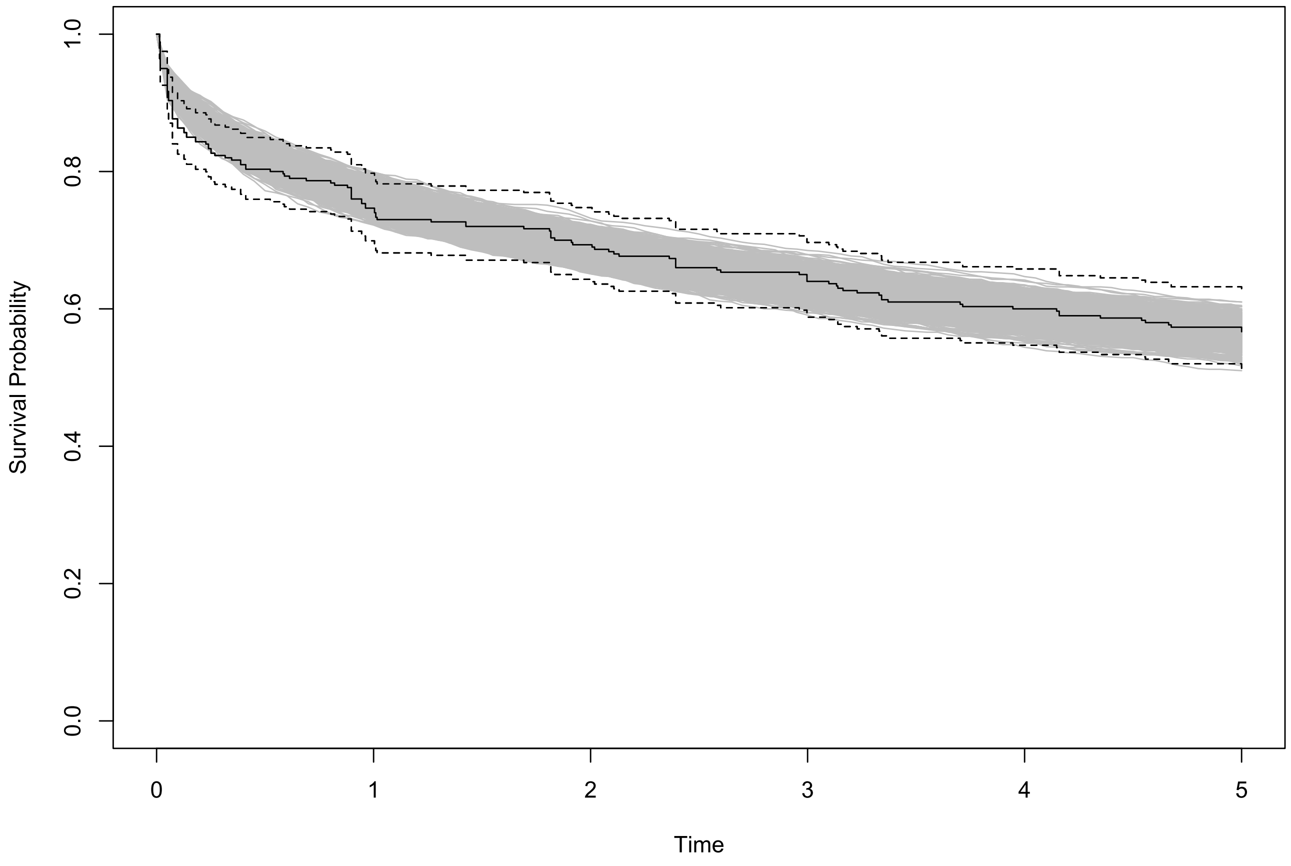

Caption: This figure shows the survival curve from Cuthbertson 2010 (13) in black (dotted lines indicate 95% confidence intervals) with simulated survival curves, from the parametric Weibull model fit, in grey. The overlap shows that the Weibull model achieves a reasonable fit to the digitally extracted data.

#### 4.4 Determining costs in the first 5 years post ICU

We combined two sources to estimate the average cost of each year of life following ICU discharge. First, we used the work of Herridge et al (23) from 2016 that calculated the annual costs of health care for a cohort of critical illness survivors who had received at least 7 days of invasive ventilation. We drew a gamma-distributed random variable for each year with means corresponding to the observed value in Herridge et al's cohort. Second, we used the data from Wodchis et al's population study of the annual costs of healthcare in Canada, which divided average costs into the costs for those from 18 to 64 years of age, and the costs for those aged 65 years and older.(24) We drew a normally distributed random variable for both age groups using the 90th percentile value observed in Wodchis et al's work to determine the mean, and then for each individual patient drew a gamma distributed random variable based on that mean and a fixed standard deviation. For survivors who were within 5 years of discharge and were invasively ventilated for at least 7 days, we took the higher of the two cost values. For all other patients, we used the Wodchis cohort costs.

#### 4.5 Incidence of eligible patients per year in Canada

The incidence of eligible patients was calculated using data from the Canadian Institute for Health Information (CIHI) gathered prior to the COVID-19 pandemic.(11) In 2013-2014 there were 4,046 patients per year classified as having respiratory failure and 5,137 patients classified as sepsis. Patients that met eligibility criteria for this study likely fell into one of those two categories (pneumonia can be classified as either). Since 2013-2014 the number of ICU admissions has likely increased (admissions in those two categories increased by 60-80% from 2007 to 2014). However, some patients classified as respiratory failure or sepsis would be invasively ventilated prior to ICU admission. In the MIMIC-IV cohort, 4.4% of all ICU admissions met the eligibility criteria for this study. In the AmsterdamUMCdb cohort, 5.5% of all ICU admissions met the eligibility criteria for this study. Given that there were more than 200,000 ICU admissions per year in Canada in 2013-2014, an estimate of 5,000 eligible patients per year in Canada is likely conservative.

#### 4.6 Probability of long-term disability in patients ventilated for less than 7 days

Prior work provides detailed information about the probability of long-term disability for patients ventilated for 7 days or more. However, for patients who are never ventilated or ventilated for less than 7 days, there is less information. Among patients ventilated for 7 days or more, those in the lowest risk group (age < 42, ICU LOS < 14 days) had a 15% probability of requiring assistance for bathing or stair locomotion at 12 months. We felt this represented an obvious upper bound. We modeled the probabilities of long-term disability in each risk group using a beta distribution. For the probability of long-term disability for patients never ventilated or ventilated for 7 days or less, we chose a beta distribution with a mean of 0.03 and a standard deviation of 0.1. This amounted to a 58% chance that the probability of long-term disability was less than 1%, and a 16% chance that the probability of long-term disability was greater than 0.1. The mean probability was 5%.

### 5 Bayesian modelling of ventilator, oxygen therapy, and ward duration

#### 5.1 Rationale

We used a cohort of non-intubated patients from the MIMIC-IV database to model the duration of ventilation (conditional on receipt of invasive ventilation), the duration of time in ICU without receiving ventilation, and the duration of a hospital ward stay after ICU discharge. This was the same cohort from which the target trial parameters were derived.(1) These were adult patients receiving oxygen via non-rebreather, high-flow nasal cannula, or non-invasive ventilation with an inspired oxygen fraction of 0.4 or higher. They had no immediate indications for invasive ventilation and no goals of care restrictions clearly prohibiting invasive ventilation.

#### 5.2 Modeling

To fit the models, we used a Bayesian accelerated failure time model for each outcome (ventilator duration, non-ventilated ICU duration, ward duration) with a Weibull hazard. Covariates included age, sex, survival (all outcomes), and invasive ventilation status (for ICU non-ventilated duration and hospital length-of-stay after ICU discharge). Note that survival could be included because in the Treeage modeling, survival was determined prior to invasive ventilation status, length of invasive ventilation, length of ICU stay, or length of hospital stay after ICU discharge. Prior distributions were weakly skeptical. Programming was done in Stan and R.(25,26) The code is available in an online repository at https://doi.org/10.5281/zenodo.7603995.

#### 5.3 Table e5 – coefficients derived from Bayesian accelerated failure time modeling

| Outcome | Coefficient | Mean | Standard deviation |
|---|---|---|---|
| Ventilator duration | intercept | 2.282046 | 0.125268 |
| | Death by day 28 | -0.45541 | 0.062491 |
| | Age | -0.25038 | 0.068236 |
| | Sex | -0.06371 | 0.036234 |
| | Log(alpha) | -0.05543 | 0.075365 |
| ICU non-ventilated duration | Intercept | 1.771127 | 0.158789 |
| | Death by day 28 | -0.42614 | 0.065718 |
| | IMV by day 28 | -0.14789 | 0.076253 |
| | Age | -0.03817 | 0.081279 |
| | Sex | 0.060461 | 0.041212 |
| | Log(alpha) | -0.15365 | 0.069422 |
| Hospital length-of-stay after first ICU discharge | Intercept | 2.121145 | 0.169543 |
| | Death by day 28 | -0.4154 | 0.072992 |
| | IMV by day 28 | -0.10609 | 0.082323 |
| | Age | -0.03265 | 0.084929 |
| | Sex | 0.083115 | 0.057048 |
| | Log(alpha) | -0.27273 | 0.078065 |

### 6 Sensitivity analysis results

#### 6.1 Table e6: Model outputs, sensitivity analysis

| | Usual care | Hypothetical threshold |
|---|---|---|
| **Durations (Days)** | | |
| ICU, non-IMV duration | 4.67 (3.34 to 6.46) | 4.72 (3.42 to 6.7) |
| IMV duration (among ventilated people) | 8.5 (5.76 to 12.4) | 8.21 (5.52 to 12.2) |
| Ward duration | 5.68 (4.24 to 7.76) | 5.73 (4.25 to 7.75) |
| **Hospital outcomes (%)** | | |
| Invasive ventilation | 27.6 (13.1 to 45.4) | 20.2 (7.6 to 38.1) |
| Survival | 74.5 (51.5 to 91.8) | 74.6 (53.5 to 92) |
| Long-term disability | 5.73 (2.1 to 10.8) | 4.56 (1.4 to 9.7) |
| **Lifetime outcomes** | | |
| Life expectancy (years) | 10.8 (7.18 to 14.1) | 10.7 (7.25 to 13.9) |
| QALYs | 8.2 (5.46 to 10.7) | 8.23 (5.58 to 10.7) |
| Cost (1000's CAD) | 74.7 (62.7 to 88.5) | 71.7 (61 to 85) |
| Net monetary benefit (1000's CAD) | 746 (474 to 990) | 751 (488 to 992) |
| **Comparative outcomes** | | |
| Probability of highest net monetary benefit | 0.497 | 0.503 |
| Incremental cost-utility ratio (CAD) | Reference | Dominant |

Caption: Model outputs (mean and 95% credible interval). The hypothetical threshold dominates usual care by providing slightly better outcomes at slightly less cost. IMV – invasive mechanical ventilation, QALYs – quality-adjusted life years, CAD = 2022 Canadian dollars. Net monetary benefit using willingness-to-pay of 100,000 CAD per QALY.

#### 6.2 Figure e2: Cost-effectiveness acceptability curve, sensitivity analysis

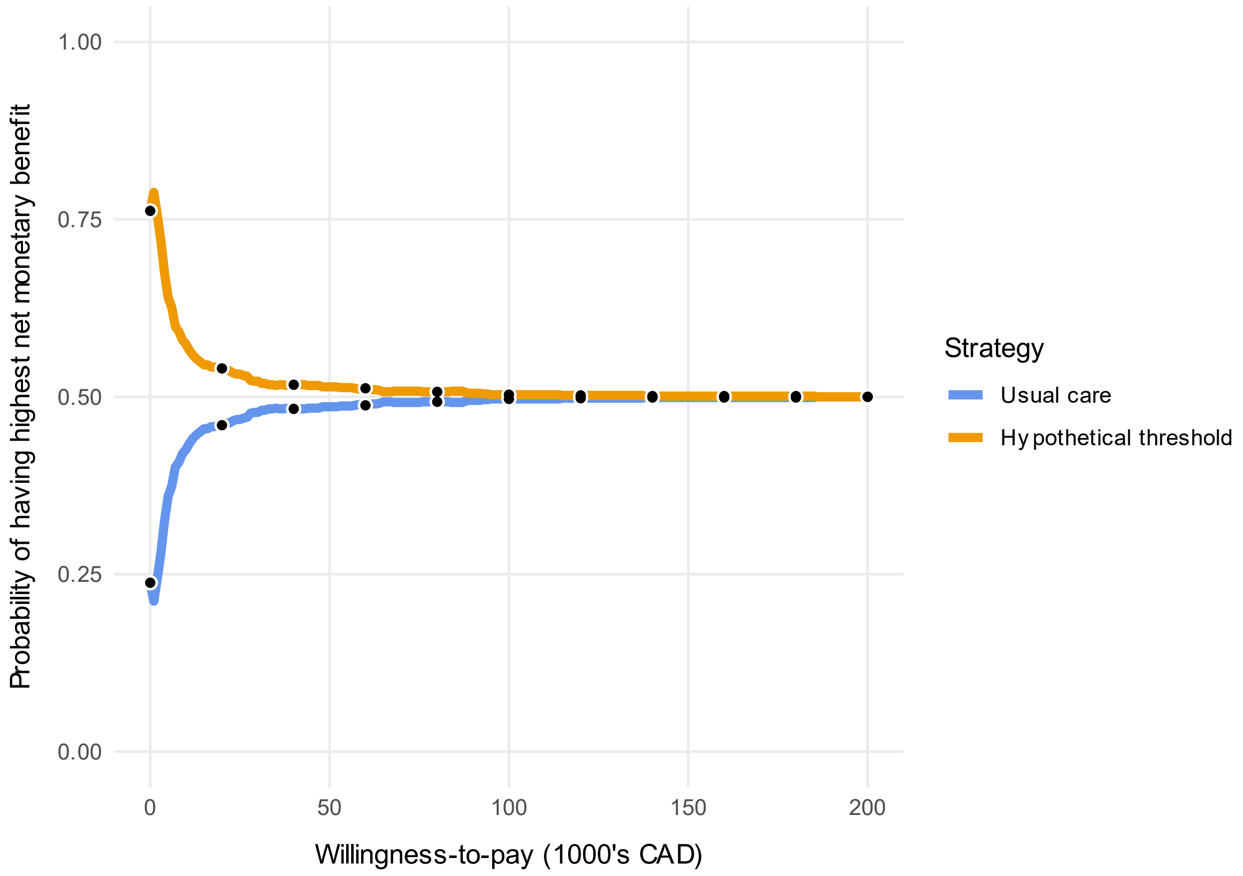

Caption: This figure shows the proportion of iterations that each strategy has the highest net monetary benefit versus willingness to pay. The strategies have equivalent net monetary benefit above a willingness-to-pay of 100,000 CAD per QALY.
